# Recommendations for Primary Prevention of Skin Melanoma

**DOI:** 10.1101/2020.08.25.20181610

**Authors:** Tõnis Tasa, Mikk Puustusmaa, Neeme Tõnisson, Berit Kolk, Peeter Padrik

## Abstract

Melanoma (MEL) is an aggressive form of skin cancer, causing over 60,000 deaths every year and it is considered one of the fastest-growing cancer forms. Genome-wide association studies have identified numerous genetic variants (SNPs) independently associated with MEL. The effects of such SNPs can be combined into a single polygenic risk score (PRS). Stratification of individuals according to PRS could be introduced to the primary prevention of melanoma. Our aim was to combine PRS with health behavior recommendations to develop a personalized recommendation for primary prevention of melanoma.

Previously published PRS models for predicting the risk of melanoma were collected from the literature. Models were validated on the UK Biobank dataset consisting of a total of 487,410 quality-controlled genotypes with 3791 prevalent and 2345 incident cases. The best performing sex-specific models were selected based on the AUC in prevalent data and independently validated on an independent UKBB incident dataset for females and males separately. The best performing model included 28 SNPs. The C-index of the best performing model in the dataset was 0.59 (0.009) and hazard ratio (HR) per unit of PRS was 1.38 (standard error of log (*HR*) = 0.03) for both males and females.

We performed absolute risk simulations on the Estonian population and developed individual risk-based clinical follow-up recommendations. Both models were able to identify individuals with more than a 2-fold risk increase. The observed 10-year risks of developing melanoma for individuals in the 99th percentile exceeded the risk of individuals in the 1^st^ percentile more than 4.5-fold.

We have developed a PRS-based recommendations pipeline for individual health behavior suggestions to support melanoma prevention.

## Introduction

Skin cancer is the most common form of cancer in Caucasians forming 40-50% of all cancer diagnoses in US (1, 2). Skin cancer can be divided into two types: non-melanoma skin cancers and melanoma. Latter being the most aggressive and lethal form of skin cancer (2). Over 280,000 new melanoma cases were diagnosed and more than 60,000 deaths were registered worldwide in 2018 (3). The number of melanoma incidences in the EU can reach up to 90,000 new cases per year, and it is considered one of the fastest-growing cancer forms in the EU (4). Melanoma is about 1.5 times more frequent in males than in females. The risk of melanoma rises with age and the average age at diagnosis is about 60 (5). Melanoma is also one of the most common types of cancer among adolescents and young adults (6). Melanoma is curable by surgical excision in the majority of cases if detected at an early stage (7).

The development of melanoma is complex, and it is interconnected with several environmental and genetic risk factors. Most cases of melanoma develop sporadically with no family history and approximately only 10% of individuals diagnosed with malignant melanoma have a first- or second-degree relative with a history of the disease (8, 9). The most significant environmental risk factor is exposure to ultraviolet (UV) radiation causing about 60-70% malignant melanomas (2, 10, 11). Solar UV radiation is 95% of UVA (315-400 nm) and it is also the primary source of radiation in tanning beds which can reach UVA doses up to 12-times that of the sun (2, 12). UVA leads to tumorigenesis primarily through oxidative stress-induced DNA damage as melanin in melanocytes acts as a photosensitizer causing diminished repair of UVA-induced oxidative damage (13, 14). However, the skin’s response to UV radiation depends on many factors including the history of tanning, hair color, and skin type (2). In addition to phenotypic and environmental factors, genetics has a role in melanoma development. Several high penetrance susceptibility genes like CDKN2A, CDK4, BRCA1, BAP1, TERT, POT1, ACD, TERF2IP, and CXC genes and some moderate penetrance susceptibility genes like MC1R and MITF are known (15). Mutation in these genes confers to a high or moderate risk of developing melanoma. However, the frequency of mutations in these genes is rare, for example, in sporadic melanoma patients without personal or familial history of melanoma the probability of mutation in the CDKN2A gene is only about 1% (16). Therefore, combining the effects of many common genomic variants to predict melanoma is reasonable.

Multiple genome-wide association studies (GWAS) have identified dozens of loci and single-nucleotide polymorphisms (SNPs) associated with melanoma (17-26). These common genomic variants can be aggregated into a single polygenic risk score (PRS). In recent years, several different PRS models have been published and It has been shown that a PRS is strongly associated with melanoma risk allowing us to stratify patients by their cancer risk (27-31). Clinical usefulness of skin cancer PRS has been analyzed by M.R. Roberts and his colleagues and they concluded that PRS might already offer meaningful improvements to risk prediction (32).

The aim of this study is to evaluate the risk prediction performance of several published melanoma PRS and to assess its use as a risk stratification approach in the context of Estonia. We aim to use information from polygenic risk stratification to propose individual follow-up actions.

## Methods

### Participant data of UK Biobank

This study used genotypes from the UK Biobank cohort (obtained 07.11.2019) and made available to Antegenes under application reference number 53602. The data was collected, genotyped using either the UK BiLEVE or Affymetrix UK Biobank Axiom Array. Melanoma cases in the UK Biobank cohort were retrieved by the status of ICD-10 codes C43. We additionally included cases with self-reported UK Biobank code “1059”.

Quality control steps and in detail methods applied in imputation data preparation have been described by the UKBB and made available at http://www.ukbiobank.ac.uk/wpcontent/uploads/2014/04/UKBiobank_genotyping_QC_documentation-web.pdf. We applied additional quality controls on autosomal chromosomes. First, we removed all variants with allele frequencies outside 0.1% and 99.9%, genotyping call rate <0.1, imputation (INFO) score <0.4 and Hardy-Weinberg equilibrium test p-value < 1E-6. Sample quality control filters were based on several pre-defined UK Biobank filters. We removed samples with excessive heterozygosity, individuals with sex chromosome aneuploidy, and excess relatives (> 10). Additionally, we only kept individuals for whom the submitted gender matched the inferred gender, and the genotyping missingness rate was below 5%.

Quality controlled samples were divided into prevalent and incident datasets for females and males separately. The prevalent datasets included MEL cases diagnosed before Biobank recruitment with 5 times as many controls without the diagnosis. Incident datasets included cases diagnosed in any of the linked databases after recruitment to the Biobank and all controls not included in the prevalent dataset. Prevalent datasets were used for identifying the best sex-specific candidate model and the incident datasets were used to obtain independent PRS effect estimates on MEL status.

### Model selection from candidate risk models

We searched the literature for PRS models in the public domain. The requirements for inclusion to the candidate set were the availability of the chromosomal location, reference and alternative allele, minor allele frequency, and an estimator for the effect size either as odds ratio (OR) or its logarithm (log-OR) specified for each genetic variant. In cases of iterative model developments on the same underlying base data, we retained chronologically newer ones. The search was performed with Google Scholar and PubMed web search engines by working through a list of articles using the search [“Polygenic risk score” or “genetic risk score” and “melanoma”], and then manually checking the results for the inclusion criteria. We additionally pruned the PRS from multi-allelic, non-autosomal, non-retrievable variants based on bioinformatics re-analysis with Illumina GSA-24v1 genotyping.

PRSs were calculated as 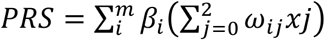, where *ω_ij_* is the probability of observing genotype *j, where j*∈*{0,1,2)* for the i-th SNP; *m* is the number of SNPs; and *β_i_* is the effect size of the i-th SNP estimated in the PRS. The mean and standard deviation of PRS in the cohort were extracted to standardize individual risk scores to Gaussian. We tested the assumption of normality with the mean of 1000 Shapiro-Wilks test replications on a random subsample of 1000 standardized PRS values.

Next, we evaluated the relationship between MEL status and standardized PRS in the two sex-specific prevalent datasets with a logistic regression model to estimate the logistic regression-based odds ratio per 1 standard deviation of PRS *(OR_sd_)*, its p-value, model Akaike information criterion (AIC) and Area Under the ROC Curve (AUC). The logistic regression model was compared to the null model using the likelihood ratio test and to estimate the Nagelkerke and McFadden pseudo-R^2^. We selected the candidate model with the highest AUC to independently assess risk stratification in the incident dataset.

### Independent performance evaluation of a polygenic risk score model

Firstly, we repeated the main analyses in the prevalent dataset. The main aim was to derive a primary risk stratification estimate, hazard ratio per 1 unit of standardized PRS (HR*_sd_*), using a right-censored and left-truncated Cox-regression survival model. The start of time interval was defined as the age of recruitment; follow-up time was fixed as the time of diagnosis. Scaled PRS was fixed as the only independent variable of MEL diagnosis status. 95% confidence intervals were created using the standard error of the log-hazard ratio.

Further, we evaluated the concordance between theoretical hazard ratio estimates derived with the continuous per unit PRS (HR_sd_) estimate and the hazard ratio estimates inferred empirically from data. For this, we binned the individuals by PRS to 5%-percentiles and estimated empiric hazard ratio of MEL directly between those classified in each bin and those within 40-60 PRS percentile. Theoretical estimates were derived from the relationship 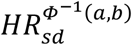, where the exponent is the expected Gaussian value between two arbitrary percentiles *a* and *b* (bounded between 0 and 1, a<b) of the Gaussian distribution, 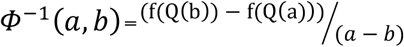, where *Q(b)* is the Gaussian quantile function on a percentile *b* and *f(Q(b))* is the Gaussian probability density function value at a quantile function value. We compared the two approaches by using the Spearman correlation coefficient and the proportion of distribution-based 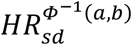 estimates in empirical confidence intervals.

### Absolute risk estimation

Individual *τ*-year (eg. 10-year) absolute risk calculations are based on the risk model developed by Pal Choudhury *et al* (33). Individual absolute risks are estimated for currently *a*-year old individuals in the presence of known risk factors (*Z*) and their relative log hazard-ratio parameters (*β*). 95% uncertainty intervals for the hazard ratio were derived using the standard error and z-statistic 95% quantiles CI_HR=exp_(*β* ±1.96*se(HR)), where se(HR) is the standard error of the log-hazard ratio estimate. Risk factors have a multiplicative effect on the base-line hazard function. The model specifies the next *τ*-year absolute risk for a currently a-year old individual as

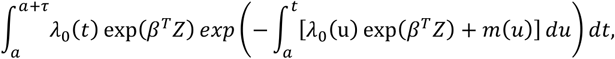

where *m(t)* is age-specific mortality rate function and *λ*_0_*(t) is* the baseline-hazard function, *t ≥ T* and *T* is the time to onset of the disease. The baseline-hazard function is derived from marginal age-specific MEL incidence rates *(λ_m_(t))* and distribution of risk factors *Z* in the general population *(F(z))*.

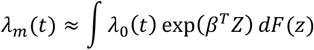

This absolute risk model allows disease background data from any country. In this analysis, we used Estonian background information. We calculated average cumulative risks using data from National Institute of Health Development of Estonia (34) that provides population average disease rates in age groups of 5-year interval. Sample sizes for each age group was acquired from Statistics Estonia for 2013-2016. Next, we assumed constant incidence rates for each year in the 5-year groups. Thus, incidence rates for each age group were calculated as *IR=X_t_/N_t_*, where X_t_ is the number of first-time cases at age *t* and N*_t_* is the total number of women in this age group. Final per-year incidences were averaged over time range 2013-2016. Age-and sex-specific mortality data were retrieved from World Health Organization (35) and competing mortality rates were constructed by subtracting yearly age- and sex-specific disease mortality rates from general mortality rates. Melanoma mortality estimates were derived from Global Cancer Observatory (36).

We applied this model to estimate absolute risks for individuals in the 1^st^, 10^th^, 25^th^, 50^th^, 75^th^, 90^th^ and 99^th^ PRS quantiles, eg. an individual on the 50^th^ percentile would have a standardized PRS of 0. Confidence intervals for the absolute risk are estimated with the upper and lower confidence intervals of the continuous per unit log-hazard ratio. Similarly, we used the absolute risk model to estimate lifetime risks (between ages 0 and 85) for the individuals in the same risk percentiles.

### Polygenic risk score based risk stratification

Next, we simulate cumulative PRS stratified lifetime risks using sex-specific MEL incidences in Estonian population by evaluating lifetime absolute risks for individuals in various PRS risk percentiles.

Our analysis first established the 10-year risk of a 50-year old female with a population average of PRS (“average individual”) using the model by Pal Choudhury (33). Additionally, we assessed the differences in ages where individuals in various PRS risk percentiles attain 1 to 3-fold increases of risk compared to the 10-year risk of an average individual of the same sex and explore the dynamics of melanoma incidences between different sexes. Lastly, we added general health and observation recommendations with special caveats for groups specifically at high risk based on current practices of melanoma monitoring.

## Results

There was a total of 487,410 quality-controlled genotypes available for males and females in the complete UK Biobank cohort. Both genotype and phenotype data were available for a total of 458 696 individuals. This included 235 578 (234 408 controls, 1170 cases) cases for incident females and 13484 (11232 controls, 2252 cases) for prevalent females, and additional 9215 prevalent (7676 controls, 1539 cases) males and 200 419 (199 233 controls, 1186 cases) incident males.

Altogether, we re-validated 5 PRS models (28, 30, 31, 37-39). We used only the PRS made up of previously known GWAS catalog SNPs and SNPs newly identified in their analysis as other compared PRS models included UKBB data in ML23 from Fritsche *et al*. (28). Normality assumption of the standardized PRS was not violated with any tested models (Shapiro-Wilks test p-values in ML23 = 0.43, ML29 = 0.50, ML33 = 0.35, ML36 = 0.13, ML37 = 0.40). The best performing model was selected based on AUC, OR_sd_, AIC and pseudo-R^2^ metrics for both females and males. The ML23 model that was based on Fritsche et al. (28) performed the best (Table 1). The model’s AUC under the ROC curve for the association between the PRS and MEL diagnosis was 0.580 (SE = 0.019) for males (Figure 1) and 0.574 (SE = 0.015) for females.

**Table 1.**
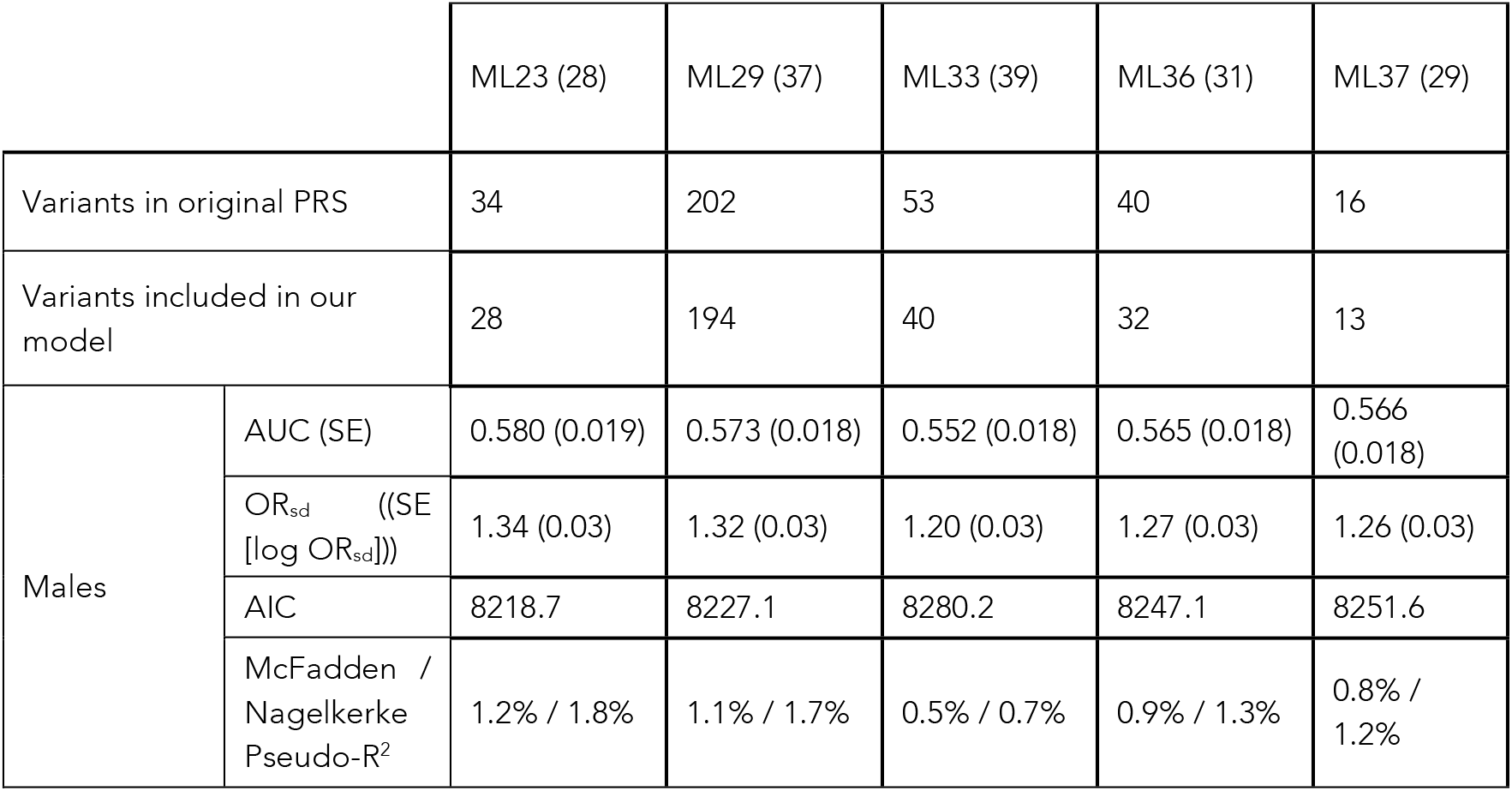
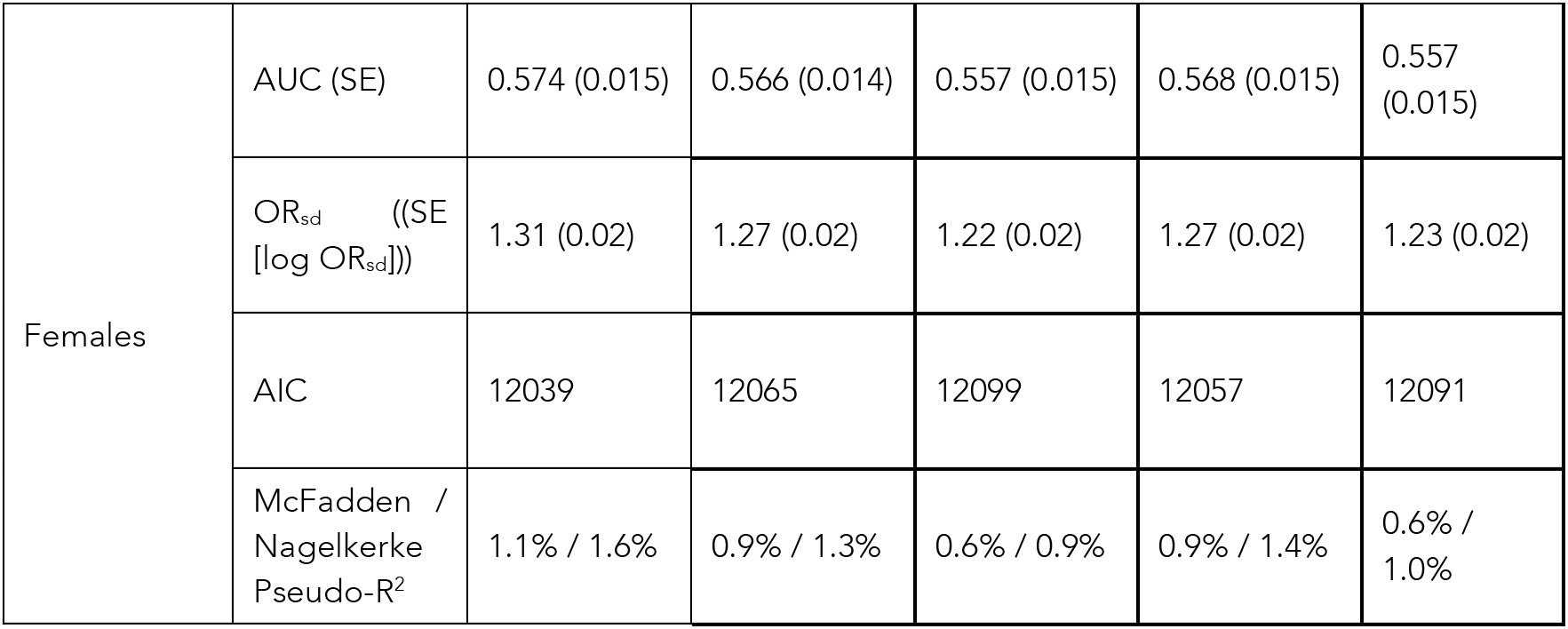
Comparison metrics of melanoma PRS models based on the prevalent UKBB dataset.

**Figure 1.**
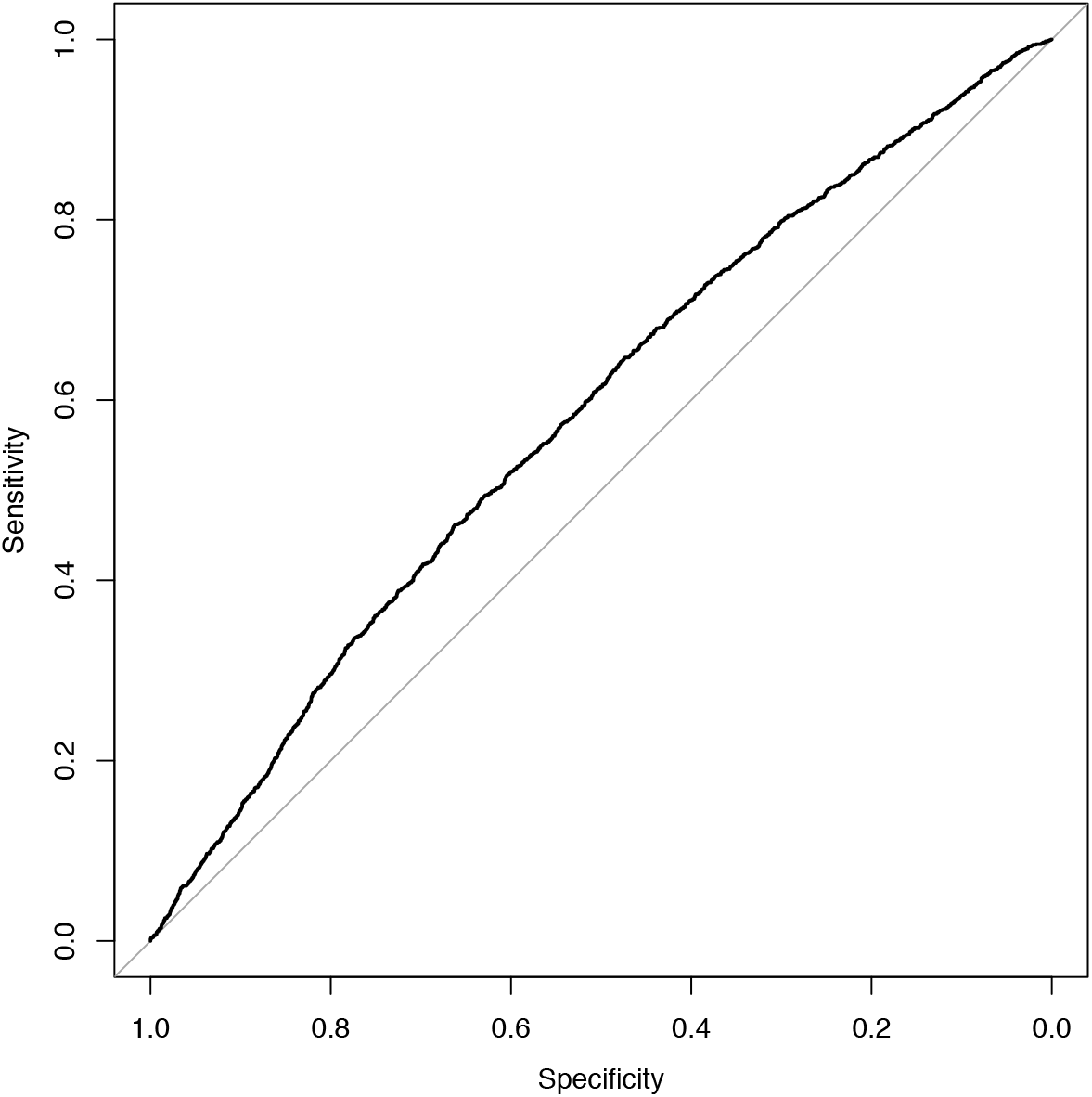
ROC plot of MEL cases and controls in UK Biobank prevalent dataset of males.

Next, we evaluated the performance of the best performing ML23 model in an independent UKBB incident dataset with the main aim of estimating the hazard ratio per unit of PRS. Table 2 presents the performance estimation metrics in the incident dataset. Hazard ratio per 1 unit of standard deviation (HR_sd_) in model ML23 was *1.38* with standard error (log *(HR))* = 0.03) for both males and females. The concordance index (C-index) of the survival model testing the relationship between PRS and MEL diagnosis status in the males’ dataset was 0.587 (0.009) and again nearly completely equivalent in females.

**Table 2.** Performance metrics of the ML23 model in the incident UK Biobank dataset.

| | | $HR_{sd}$ (95% confidence interval) | C-index (SE) | -2 x log likelihood | Likelihood ratio test p-value |
| --- | --- | --- | --- | --- | --- |
| ML23 (5) | Males | 1.38 (1.31-1.47) | 0.587 (0.009) | 120.3 | < 2e-16 |
|  | Females | 1.38 (1.3 – 1.46) | 0.588 (0.009) | 117.1 | < 2e-16 |

Hazard ratio estimates compared to individuals in 40-60 percentile of PRS are visualized in Figure 2. In females’ dataset (panel B), the theoretical hazard ratio matched in empirical estimate’s confidence intervals in 15 out of 16 comparisons (Spearman correlation 0.973). In males’ dataset (panel A), the intervals matched in all comparisons (Spearman correlation 0.948).

**Figure 2.**
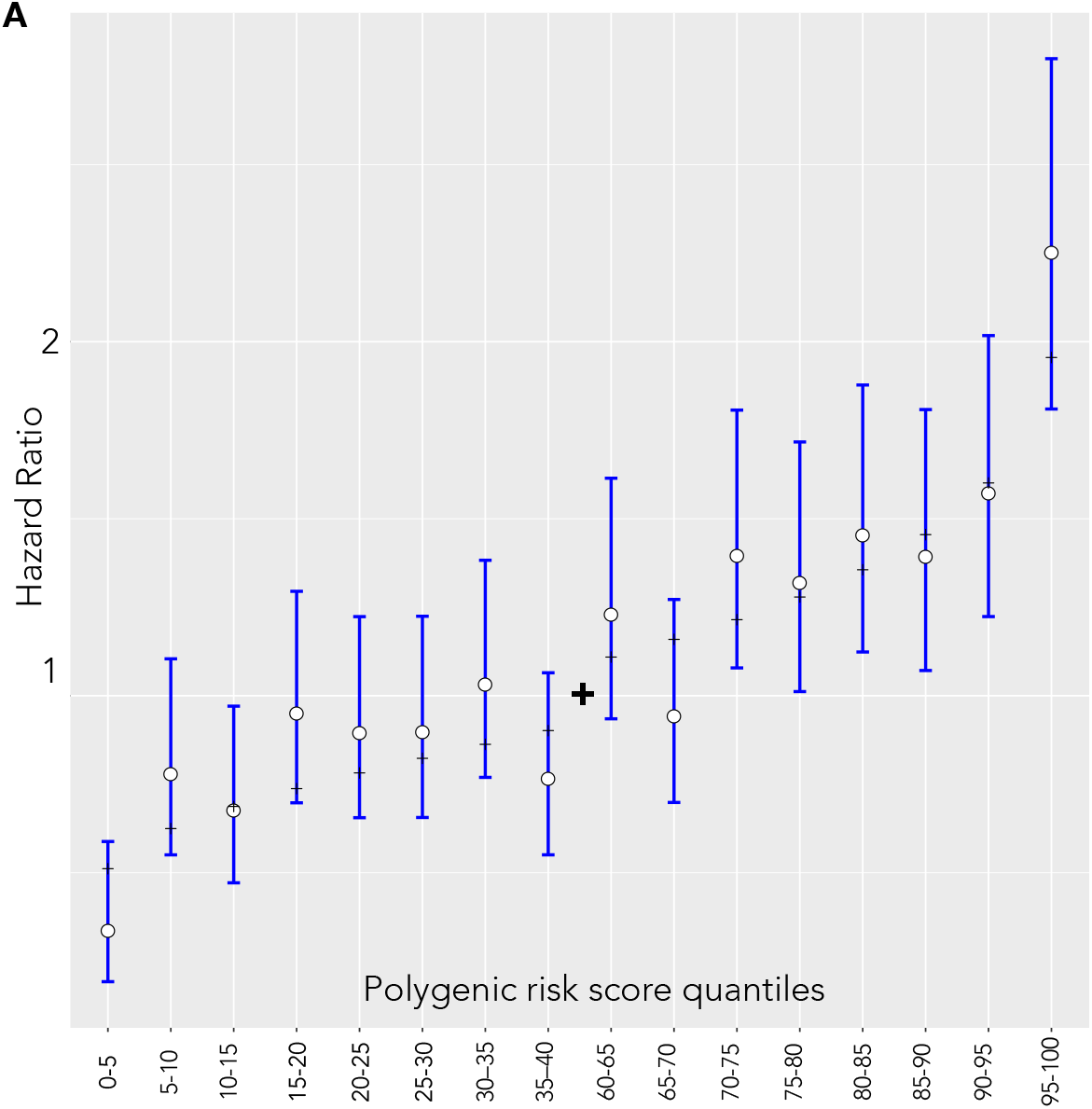
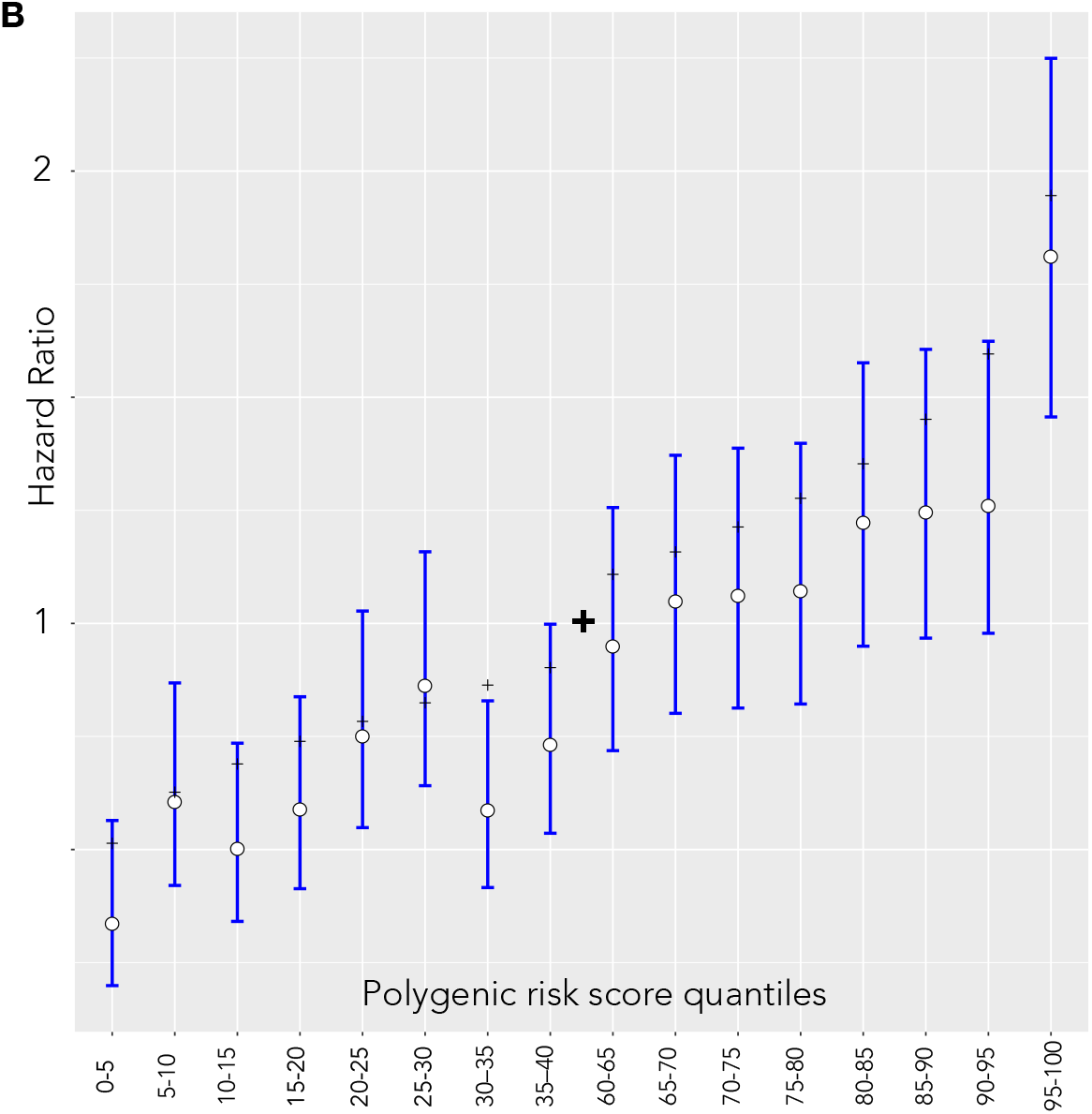
Hazard ratio estimates between quantiles 40-60 of the PRS and categorized 5% bins in the UKBB incident dataset. White dots and blue lines represent empirically estimated hazard ratio estimates and corresponding confidence intervals. Black dashes represent the theoretical hazard ratio for the 5%-quantile bins derived from the hazard ratio of per unit PRS. (A) Males (B) Females

### Risk stratification with a melanoma polygenic risk score

We used a model by Pal Choudhury et al. (33) to derive individual 10-year risks and specified *F(z)* as the distribution of PRS estimates in the whole UKBB cohort. The log-hazard ratio (*β*) is based on the sex-specific estimate of the log-hazard ratio in the ML23 model of the incident UKBB dataset. Age-specific MEL incidence and competing mortality rates provided the background for MEL incidences in the Estonian population.

In the Estonian population, the absolute risk of developing melanoma in the next 10 years among 50-year old men in the 1^st^ percentile is 0.08% (0.07% – 0.10%) and 0.37% (0.33% – 0.42%) in the 99^th^ percentile (0.12% (0.10% – 0.14%) and 0.53% (0.47% – 0.60%) for women, respectively). At age 70, corresponding risks for the same percentiles of men become 0.20% (0.18% – 0.24%) and 0.93% (0.83% – 1.04%). As opposed younger ages, at age 70 the 10-year risk of females is lower than of males: 0.17% (0.14% – 0.20%) and 0.75% (0.67% – 0.84%) respectively in the most extreme percentiles. The relative risks between the most extreme percentiles are therefore around 4.4-fold. Competing risks accounted cumulative risks of females in the 99th percentile surpass 2.13% (1.89% – 2.40%) by age 85 but remain at 0.47% (0.41% – 0.55%) in the 1^st^ percentile (Figure 3). Equivalent values for women in Estonia exceed those of men: 2.74% (2.44% – 3.07%) and 0.62% (0.53% – 0.72%) in the most extreme percentiles.

**Figure 3.**
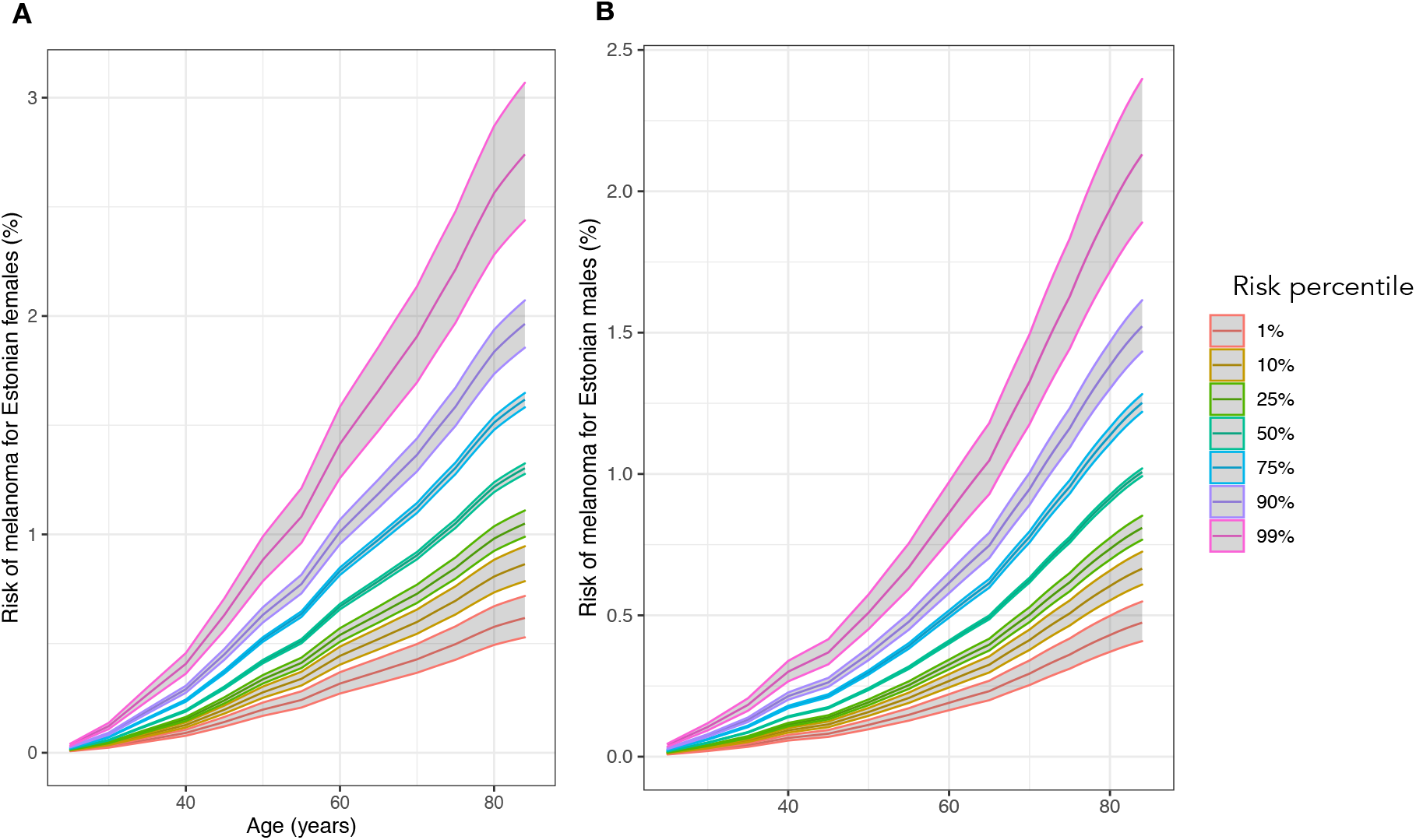
Cumulative risks (%) of melanoma between ages 20 to 85 in various risk percentiles. (A) Females (B) Males

A genetically average 40-year old has a 10-year risk of melanoma equaling 0.1% in males (Figure 4 panel A) and 0.218% in females (Figure 4 panel B). A 30-year-old male in the 99^th^ percentile (26 for females) has a larger risk than an average 40-year-old. At the same time, males in the 1^st^ percentile attain this risk by age 54. Females in the lowest 5% do not attain the average 40-year old’s absolute average risk even by age 70. More than 1% of males and females have a 2-fold risk compared to average at age 40. The 10-year risk of a 40-year-old male doubles by age 56 in genetically average males and triples by age 63. Importantly, such risk increases do not occur in females that have a larger proportion of incidences in younger ages. The 1^st^ percentile of males only triples the population average 40-year-old’s risk by age 70. This does not occur in females.

**Figure 4.**
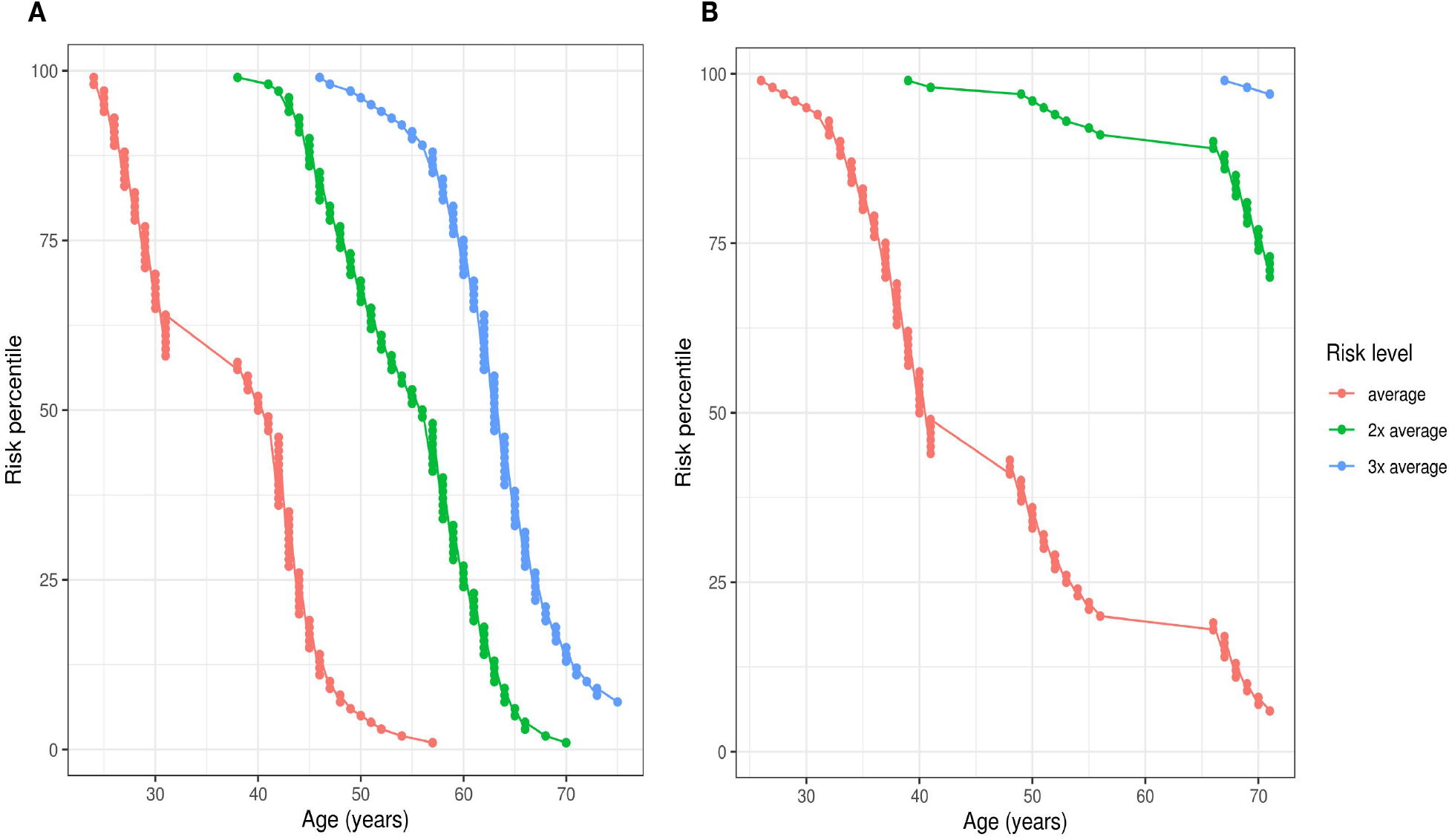
Ages when Estonian individuals in different risk percentiles attain 1-3 fold multiples of 10-year risk compared to 40-year olds with population average PRS (Risk level: “average”). (A) Males (B) Females,

No population screening approaches have been instituted to track and screen melanoma. Early detection can be supported with regular self-examinations and health behavior that apply to all. Also, we recommend once-a-year dermatology visits with individuals having greater than 2-fold risk increases compared to those with average PRS.

1. Recommendations for all
  a. Watch for abnormal moles and perform a skin self-exam every month for any new changes. If you spot anything that just does not look right, get it checked by a dermatologist as soon as possible.
  b. Limit your exposure to ultraviolet (UV) radiation: protect your skin by using a broad-spectrum sunscreen with an SPF 30 or higher
  c. Avoid using tanning beds and sunlamps.
2. Relative risk increases over 2-fold
  a. See a dermatologist once a year for a full-body skin exam.

## Discussion

A record number of new skin cancers cases are seen worldwide causing a significant burden to the health care system (2, 40). Melanoma is the most lethal form of skin cancer and in the U.S., the cost of melanoma treatment is about $3.3 billion each year (2, 41). The risk of melanoma rises with age and the average age at diagnosis is about 60. With an aging population, the number of melanoma cases will likely remain high for the near future. It has been shown that genetics has a role in melanoma development. Common genomic variants can help identify people at high risk for melanoma, particularly those who lack traditional risk factors (31).

In this study, we validated different publicly available PRS models to find a sex-specific model that could be used to design a novel risk-based screening strategy for melanoma. Our best-performing model, named ML23, was a pruned version of Fritsche et al (28) PRS model containing a total of 28 SNPs. In the UK Biobank incident dataset, the AUC was 0.58 and the estimated hazard ratio (HR) for overall MEL per unit PRS was 1.38 for both sexes. Our model was able to identify individuals with more than 4-fold PRS-based risk differences between the extremes.

We also developed personalized health behavior recommendations based on PRS result and overall guidelines for melanoma prevention. First, as nevi are one of the strongest risk factors for melanoma (42) we recommend to watch for abnormal moles and perform a skin self-exam every month for any new changes. Second, it is estimated that 60-70% of melanomas are caused by ultraviolet (UV) radiation (2, 11). Thus, primary prevention strategies that reduce sun exposure and encourage increased sun protection are important for reducing melanoma incidence and mortality (10, 43-45). Therefore, we recommend to limit exposure to ultraviolet to all patients and suggest protecting their skin by using a broad-spectrum sunscreen with an SPF 30 or higher. Besides, as tanning beds can reach UVA doses 12-times that of the sun (12) we recommend all patents to avoid using tanning beds and sunlamps if possible, regardless of the fact that skin’s response to UV radiation depends on many factors including history of tanning, hair color, skin type and genetic background (2). For individuals, whose relative risk is increased over 2-folds, we recommend full-body skin exam by a dermatologist once a year. Survival from melanoma is strongly related to tumor thickness and there is strongest evidence that whole-body skin examination reduces the incidence of thick melanoma (43). Our approach is easily adaptable to other nationalities by using population background information data of other genetically similar populations.

In conclusion, our PRS based model identifies individuals at more than 2-fold risk and provides recommendations for melanoma prevention.

## Data Availability

Individual level genotype and phenotype data from UK Biobank can not be explicitly shared. The UK Biobank Resource was used under Application Reference Number 53602. New users can request access to UK Biobank from http://www.ukbiobank.ac.uk/resources/.

## Acknowledgments

This research has been conducted using the UK Biobank Resource under Application Reference Number 53602 and with support by EIT Health The Digital Sandbox program.

## Ethical approval

UK Biobank: The UK Biobank study was approved by the North West Multi-Centre Research Ethics Committee (UK Biobank reference: 16/NW/0274). All participants provided written informed consent to participate in the UK Biobank study.

## Funding

OÜ Antegenes has received a grant from the EIT Health The Digital Sandbox program and additional Innovation Voucher funding meant for business development of small and medium sized Estonian enterprises.

## Notes

### Competing Interest Statement

The authors have declared no competing interest.

### Funding Statement

OU Antegenes has received a grant from the EIT Health The Digital Sandbox program and additional Innovation Voucher funding meant for business development of small and medium sized Estonian enterprises.

